# inClinico Agent for clinical trial endpoint prediction and prospective evaluation

**DOI:** 10.64898/2026.10.13.26365373

**Authors:** Jianjiu Chen, Chengchen Duan, Wenhao Gou, Jingwan Wang, Zetian Zheng, Howell Leung, Vladimir Naumov, Denis Sidorenko, Alex Aliper, Feng Ren, Frank W. Pun, Alex Zhavoronkov

## Abstract

High attrition rates across clinical drug development impose immense clinical and financial burdens, yet prospectively forecasting whether an individual trial will achieve its primary endpoint remains challenging. Here we present a machine-learning system that predicts clinical trial success across phases I-IV, independently modeling studies with efficacy versus safety/toxicity primary endpoints. To prevent lookahead bias and outcome leakage, models are trained on registration-time features, including target and mechanism-of-action precedent, drug modality and novelty, trial design and operational characteristics, and frozen literature-derived target-choice embeddings. Each prediction yields a continuous Predictive Trial Index (PTI) between 0 and 1 reflecting success probability. On a test set of trials initiated in 2021 or later, the efficacy model achieved an area under the receiver operating characteristic curve (AUROC) of 0.828 (95% CI 0.812–0.845; n = 2,347) and the safety/toxicity model reached 0.809 (95% CI 0.787–0.831; n = 1,485). These models are embedded in inClinico Agent, an agentic AI system integrating frontier large language models (LLMs) with deterministic scoring engines, retrieval tools and specialized clinical skills. We also introduce ClinicoBench, a prospective benchmark of 3,235 ongoing trials evaluated by inClinico Agent and five frontier LLMs, establishing a leakage-free foundation for tracking prospective predictive accuracy as trials reach completion.

## Introduction

A large proportion of clinical trials fail, and it is difficult to predict in advance whether any individual trial will succeed or fail. Across the full development pipeline, fewer than one in seven programs that enter clinical testing ultimately reach approval, and attrition is distributed across every phase (Wong et al. 2019). Because each failure consumes years of effort and substantial capital, and because the decision of which trials to run is still made largely on the basis of expert judgment — a process that is hard to reproduce and harder to audit — a reliable, quantitative estimate of a trial’s probability of success would be of considerable value to the field.

To address this gap, the original inClinico combined a target-choice model with a trial-design model and reported an AUROC of 0.88 for the Phase II-to-III transition in a quasi-prospective evaluation, with a genuinely prospective series of 17 trials that later read out (Aliper et al. 2023). By design, however, it addressed only this single transition. The present work extends that line, predicting primary-endpoint attainment across all phases and for both efficacy and safety/toxicity endpoints and evaluating these predictions prospectively on a large set of ongoing trials.

Other approaches include HINT, which integrates multimodal trial, drug and disease information, and SPOT, which models relations among trials over time (Fu et al. 2022; Wang et al. 2023). Public resources such as TrialBench and the Clinical Trial Outcome database, and models such as LIFTED, broaden the available tasks, datasets and modeling strategies (Chen et al. 2025; Gao et al. 2026; Zheng et al. 2025). Furthermore, Jin et al. (2024) evaluate general-purpose LLMs directly on trial-outcome prediction. Agent-based systems including ClinicalAgent and AutoCT combine clinical-trial reasoning or feature generation with predictive tools (Yue et al. 2024; Liu et al. 2025), and CT Open provides a complementary live forecasting challenge with time-stamped evaluation and screening for public outcome disclosure (Wang et al. 2026). inClinico Agent and ClinicoBench build on these efforts, pairing endpoint-specific prediction with an agentic analysis system and a prospective, continuously updated benchmark.

Here we develop two models to predict primary-endpoint attainment, one for trials classified as having efficacy endpoints and one for trials classified as having safety/toxicity endpoints, using records spanning Phases I–IV. The models draw on drug target, mechanism of action (MOA), modality, trial design and operational characteristics, together with representations of the target–disease relationship derived from a biomedical language model. Each trial receives a Predictive Trial Index (PTI) between 0 and 1 that scales with the model’s estimated likelihood that the trial will meet its primary endpoint.

The predictive models form the quantitative core of inClinico Agent, an agentic AI system for clinical-trial analysis. The system translates natural-language queries into workflows that combine LLMs with tools for retrieval, scoring and figure generation. Figure 1 shows the architecture: a routing layer selects skills and tools; agents plan, execute, critique and report; and the critique step can return a task to planning. Internal and external databases, clinical-analysis skills and a session workspace support these operations. In the following sections, we first develop and evaluate the predictive models that anchor the system, and then return to the agent and its skills in the Discussion.

**Figure 1.**
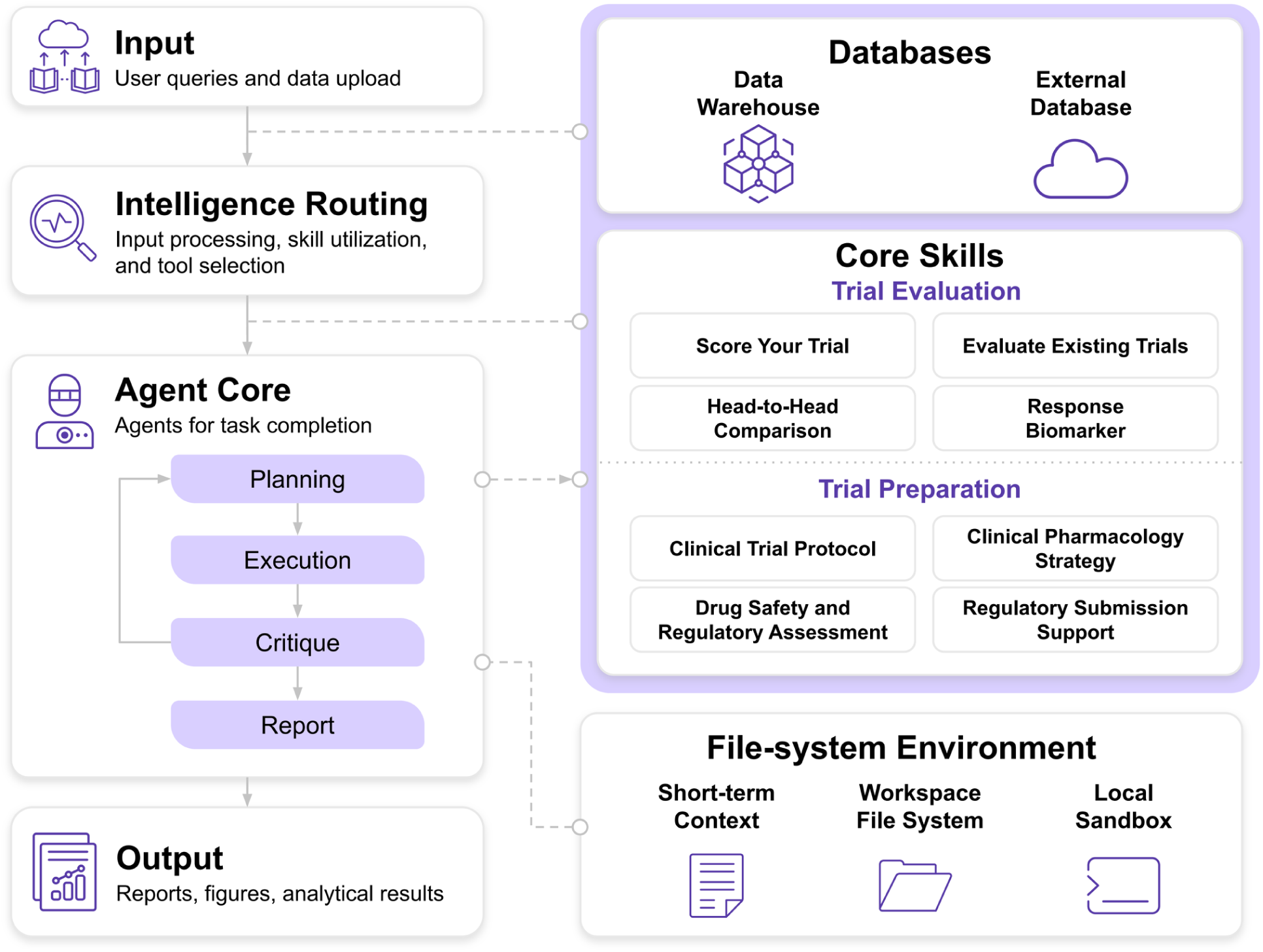
Architecture of inClinico Agent. The left column traces a request from input to output. User queries and uploaded data enter at the Input layer. The Intelligence Routing layer processes the input and selects the skills and tools required for the task. The Agent Core contains agents that complete the task in four sequential stages: Planning, Execution, Critique, and Report. A feedback loop returns the result of the Critique stage to the Planning stage. The Output layer delivers reports, figures, and analytical results. The right column shows the resources available to the agents. The shaded block groups the Databases and the Core Skills. The Databases comprise an internal data warehouse and external databases. The Core Skills are skill sets for clinical trial analysis, grouped into Trial Evaluation and Trial Preparation. The File-system Environment provides a file-based workspace and a secure execution sandbox for long-running analyses. Its three components are short-term context, the workspace file system, and a local sandbox. Dashed connectors mark where the left column draws on these resources: the Databases between Input and Intelligence Routing, the Core Skills between Intelligence Routing and Agent Core and from the Planning stage, and the File-system Environment from the Critique stage.

## Methods

### Curated clinical trial corpus

inClinico Agent is built on a database of over 102,618 curated clinical trial records, which supports real-time semantic querying across drug, target, indication, and trial phase. Every trial in this database has a resolved drug target and disease and carries either efficacy or safety/toxicity as its primary endpoint. Each record integrates structured metadata, including drug names, therapeutic modality, target mechanisms, indication, design characteristics, and trial outcomes. For modeling, each trial is assigned a binary outcome based on its reported outcomes: a trial that meets its primary endpoints is labeled a success, and a trial that does not meet its primary endpoints is labeled a failure. Each trial is then routed to the efficacy model or the safety/toxicity model based on the type of primary endpoint the sponsor registered; when a trial lists both an efficacy and a safety/toxicity endpoint as primary, it is assigned according to its first-listed primary endpoint. The corpus is continuously refreshed through a data pipeline that combines human expert and LLM-assisted curation.

### Feature engineering

The feature set was shaped by three considerations: the availability of each field in the curated trial metadata; discussion with clinical-trial domain experts about which biological, design, and operational factors plausibly bear on a trial’s outcome; and iterative experimentation on the training and validation data. Every feature is computed only from information available at the time of trial design. Following the structure summarized in Figure 2, the features are organized into three groups: (I) Biology Evidence, (II) Trial Design, and (III) Trial Operation. In total, the efficacy model uses 44 features and the safety/toxicity model 41.

**Figure 2.**
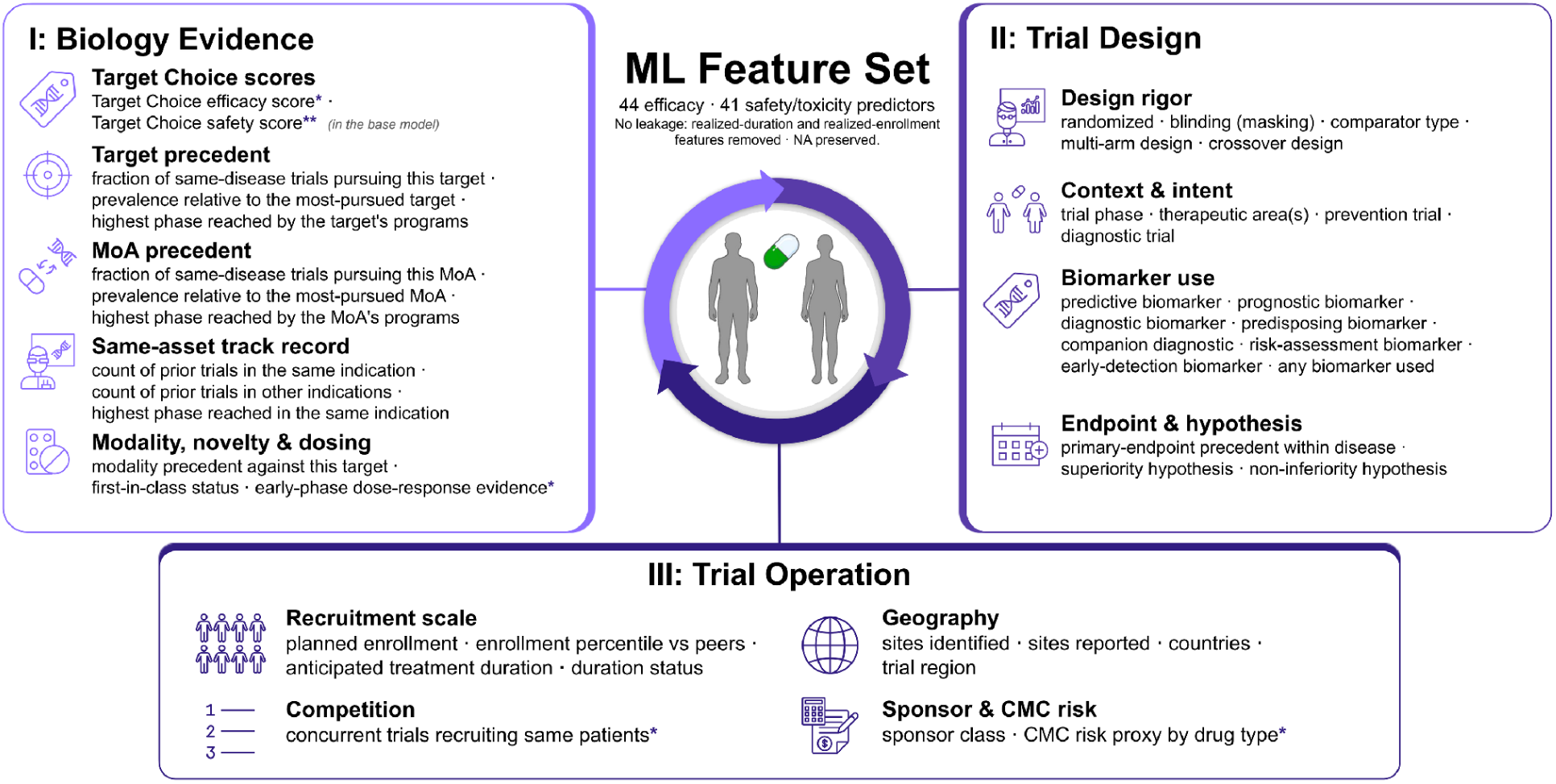
Registration-time feature set of the endpoint-specific machine learning (ML) models. All features are computed from information available at the time of trial design. The features are organized into three groups: (I) Biology Evidence, (II) Trial Design, and (III) Trial Operation. Within each group the feature families are named in bold above their individual features. The efficacy model uses 44 features, and the safety/toxicity model uses 41. A single asterisk (*) marks features used only by the efficacy model: Target Choice efficacy score, early-phase dose-response evidence, concurrent trials recruiting the same patients, and CMC risk proxy by drug type. A double asterisk (**) marks the Target Choice safety score, which only the safety/toxicity model uses. All other features feed both models. CMC, chemistry, manufacturing, and controls; MoA, mechanism of action; NA, not available.

### Group I: Biology Evidence

This group captures what is already known about the target, mechanism, asset, and modality. It includes target-choice efficacy and safety scores (detailed in the next subsection); *target precedent* features — the fraction of same-disease trials pursuing the target, the target’s prevalence relative to the most-pursued target in the disease, and the highest phase the target’s programs have reached; the analogous *MOA precedent* features for the mechanism of action; a *same-asset track record* — the counts of prior trials of the same asset in the same and in other indications, and the highest phase the asset has reached in the same indication; and *modality, novelty and dosing* features, including the prevalence of the drug modality against the target, first-in-class status, and, for the efficacy model, whether an earlier same-drug, same-disease study showed a dose-response signal.

### Target-choice efficacy and safety scores

Two of the Biology Evidence features — a target-choice efficacy score and a target-choice safety score — summarize, from the target and indication alone, how promising that biological choice has historically been. They are produced by a three-stage pipeline. First, the trial’s disease and primary-drug-target free-text fields are each encoded with a frozen biomedical language model — a BioBERT checkpoint whose pre-training is fixed to 2019 (Lee et al. 2020) — and the two representations are concatenated into a single target-plus-disease embedding. Because the encoder is never fine-tuned on trial outcomes and its knowledge predates the 2021-and-later test trials, it cannot encode the result of any trial in the test set.

Second, a supervised head is trained on these embeddings to predict trial success, with a separate head for each endpoint family — one trained on efficacy-labeled trials (yielding the target-choice efficacy score) and one on safety/toxicity-labeled trials (yielding the target-choice safety score). For each head we tuned five candidate learners — logistic regression, a multilayer perceptron, XGBoost, LightGBM, and CatBoost — by time-blocked cross-validation on the training trials (start years 2000–2020), and selected the best-performing learner by its discrimination on the test trials (start year 2021 or later); XGBoost was selected for both endpoints. Because the heads are never trained on the test trials, and the encoder predates them, the target-choice scores introduce no leakage onto the test set.

Third, we must decide what score to assign each training trial when the score is subsequently used as a feature by the downstream endpoint model. For the training trials, each score is produced by random five-fold cross-fitting of the selected head, so that a trial is always scored by a copy of the head that was not fit on that trial; test and future trials, which the head never saw, simply receive the full-model score. One limitation of this scheme is that the cross-fitting is not time-blocked, so when scoring an earlier trial a fold’s head may have been fit on later trials — a minor, acknowledged source of look-ahead confined to the training set. We nonetheless prefer cross-fitting to the alternative of scoring every training trial with a single head fit on the entire training set, because such in-sample scores would be optimistically high and would lead the downstream model to over-rely on this one feature. The cross-fitted training scores are not inflated: their discrimination on the training trials (AUROC about 0.72 for efficacy and 0.64 for safety/toxicity) is no higher than the selected head’s discrimination on the test trials (about 0.73 and 0.76), confirming that the scores entering the downstream model reflect genuine out-of-sample signal rather than memorized labels. The downstream model therefore treats the target-choice score as one informative feature among many rather than relying on it disproportionately.

### Group II: Trial Design

This group encodes the rigor and intent of the protocol: design-rigor features (randomization, masking, comparator type, multiple-arm and crossover design); context-and-intent features (trial phase, therapeutic area, and whether the trial is a prevention or diagnostic study); biomarker-use features (predictive, prognostic, diagnostic, predisposing, companion-diagnostic, risk-assessment, and early-detection biomarkers, and whether any biomarker is used); and endpoint-and-hypothesis features (the prevalence of the trial’s primary-endpoint type within the disease, and whether the design states a superiority or a non-inferiority hypothesis).

### Group III: Trial Operation

This group describes the scale and footprint of the study: recruitment scale (planned target enrollment and its percentile against peer trials in the same disease and phase band); geography (numbers of identified and reported sites, number of countries, and trial region); competition; and sponsor and manufacturing risk. The recruitment-competition feature (used by the efficacy model) is the count of other trials competing for the same patient pool, defined as trials that share at least one indication token and at least one country with the index trial and that began within a twelve-month window around it. The CMC-risk proxy by drug type feature (used by the efficacy model) is an ordinal chemistry-manufacturing-and-controls risk score, assigned from the trial’s drug modality by a rule-based hierarchy that ranges from low risk for small molecules and simple peptides, through moderate risk for standard biologics, to high and extreme risk for engineered biologics, nucleic-acid therapeutics, and cell and gene therapies; when a trial lists several modalities, the riskiest component determines the score. Two features illustrate how the design-time constraint is enforced in practice. Recruitment scale uses the planned target enrollment specified at registration, not the actual enrollment achieved, which is unknown at trial design; and the treatment-duration feature uses only the treatment duration specified in the study design, not the actual treatment duration, which becomes known only once the trial ends.

### Model training

Before modeling, we excluded trials that were still ongoing and trials that had completed but whose outcome was unknown, retaining only trials with a resolved success or failure label. The remaining trials were split into training and test sets by start year (Figure 3). The training set comprises trials with a start year from 2000 through 2020; we use 2000 as the lower bound because ClinicalTrials.gov, an important public registry underlying the corpus, launched in 2000. The test set comprises trials with a start year of 2021 or later and was reserved for evaluation. This yielded 28,265 trials for training and time-separated validation and 3,832 trials for testing.

**Figure 3.**
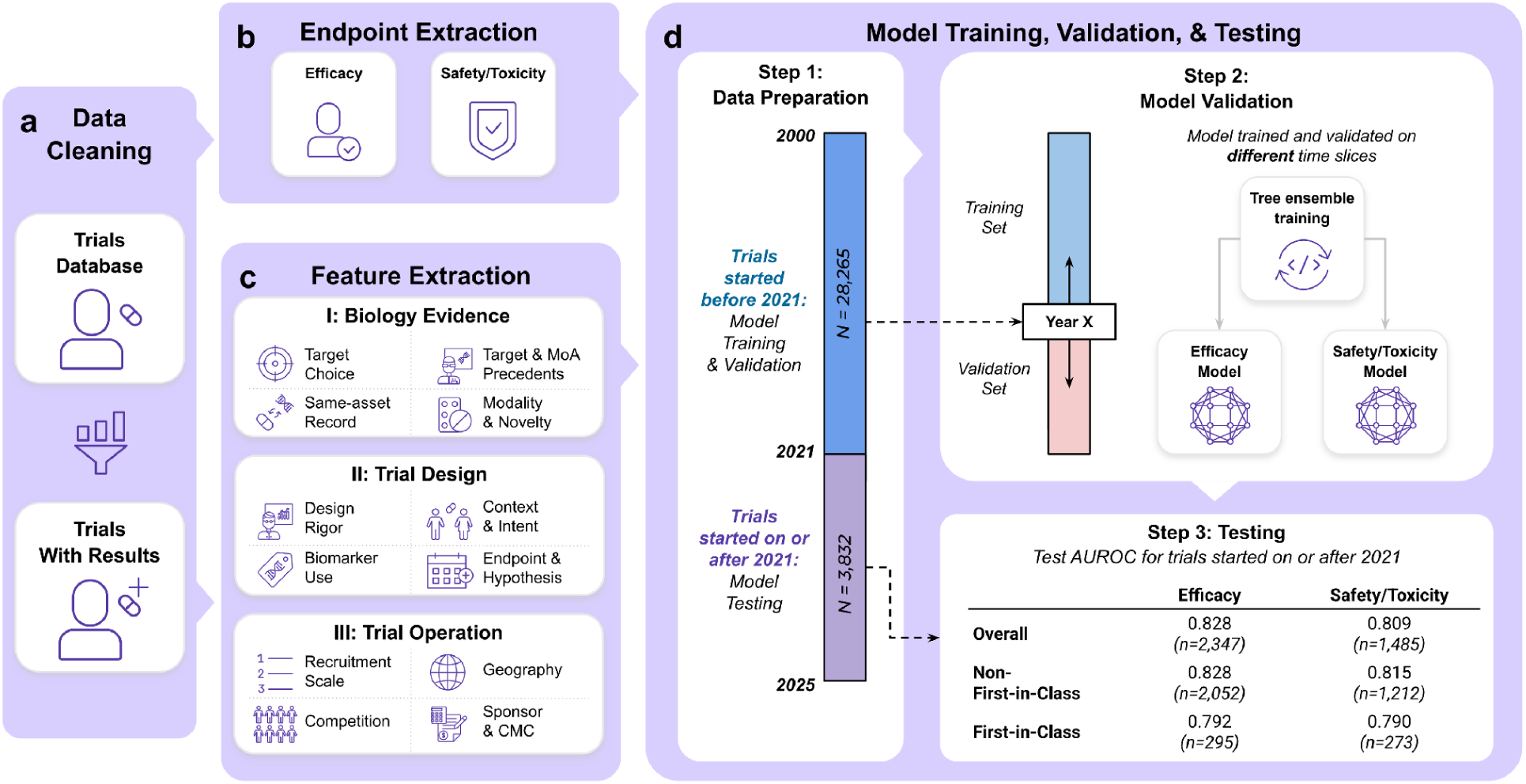
Workflow for model training, validation, and testing of the endpoint-specific models. **a)** Data cleaning of the trials database retains trials with results, defined as trials with a resolved success or failure label. **b)** Endpoint extraction assigns each trial an efficacy or safety/toxicity endpoint label. **c)** Feature extraction assigns each trial a registration-time feature set in three groups. Group I (Biology Evidence) comprises target choice, target and MoA precedents, same-asset record, and modality and novelty. Group II (Trial Design) comprises design rigor, context and intent, biomarker use, and endpoint and hypothesis. Group III (Trial Operation) comprises recruitment scale, geography, competition, and sponsor and CMC. **d)** Model training, validation, and testing. Step 1 (data preparation) assigns trials started from 2000 through 2020 (N = 28,265) to training and time-separated validation. Trials started in 2021 or later (N = 3,832) form the test set. Step 2 (model validation) selects hyperparameters by time-blocked cross-validation, in which each validation fold occurs after its training data (Year X marks the split point). Tree-ensemble models (XGBoost, LightGBM, and CatBoost) are then trained for the efficacy endpoint and the safety/toxicity endpoint. Step 3 (testing) reports the test AUROC by endpoint family for all test trials and for non-first-in-class and first-in-class trials. AUROC, area under the receiver operating characteristic curve; CMC, chemistry, manufacturing, and controls; MoA, mechanism of action; N, number of trials in each period; n, number of test trials in each cell.

The two endpoint families were modeled independently. For each family we compared four frameworks — logistic regression, XGBoost (Chen and Guestrin 2016), LightGBM (Ke et al. 2017), and CatBoost (Prokhorenkova et al. 2018); the comparison and the resulting choice of framework are reported in Results (Model selection). Each framework was given the preprocessing it requires. For the gradient-boosted tree ensembles (XGBoost, LightGBM, and CatBoost), numeric features are passed through with missing values preserved and handled natively, and categorical features are encoded by framework: one-hot indicator columns with a dedicated missing-value category for XGBoost, and native categorical handling for LightGBM and CatBoost (with CatBoost representing a missing value as an explicit category). Logistic regression, which cannot accept missing values and is sensitive to feature scale, is fit as a pipeline that median-imputes missing values and then standardizes every feature — with the imputation medians and the standardization mean and variance learned on the training fold only — and uses the same one-hot categorical encoding as XGBoost.

Within the training set, hyperparameters were selected by time-blocked cross-validation. Trials were ordered by start year and partitioned into five expanding-window folds with scikit-learn’s TimeSeriesSplit, so that each validation fold is a contiguous block of later-starting trials and the trials used to fit a fold always precede it in time, mirroring the deployment setting in which the model must predict the future from the past; the five validation folds are successive, approximately equal-sized blocks of trials, and the same partition — determined by the start-year ordering — is used for every framework. Hyperparameters were searched with Optuna using a tree-structured Parzen estimator sampler (Akiba et al. 2019), run for 40 trials per model and scored by the mean validation ROC-AUC across the five folds. The search spaces covered the standard hyperparameters of each framework: for the tree ensembles, tree depth or number of leaves, learning rate, row and column subsampling, and L1/L2 regularization, with the number of boosting rounds set on each fold by early stopping; and for logistic regression, the inverse regularization strength. After the best configuration was selected, each model was refit on the entire training set and applied once to the test set.

We report 95% confidence intervals using DeLong’s analytic variance for AUROC (DeLong et al. 1988), Wilson intervals for accuracy, precision and recall (Wilson 1927), a normal approximation combining sensitivity and specificity variances for balanced accuracy, and an outcome-stratified bootstrap with 2,000 replicates for AUPRC and F1 (Efron and Tibshirani 1994). These intervals treat trials as independent; to the extent that some assets or development programs contribute several correlated trials, the effective sample size is smaller than the trial count and the intervals may modestly understate uncertainty. We did not adopt a correlation-aware interval, such as a cluster bootstrap grouped by asset, judging the added complexity unwarranted given that this effect is expected to be small.

### ClinicoBench: prospective benchmarking against LLMs

To evaluate predictive ability prospectively and against general-purpose reasoning systems, we assembled ClinicoBench, a benchmark of 3,235 ongoing Phase I–III clinical trials. Each benchmarked trial has either efficacy or safety as its registered primary endpoint, has not yet released results, and is expected to complete on or before 31 December 2027. Because we verified that all included trials are either actively recruiting or active but no longer recruiting, all trial outcomes are, by construction, unknown at the time of prediction. We compared inClinico Agent against five frontier large language models — DeepSeek-V4-Pro, GPT-6-Astra, Grok-4.6, Claude-Fable-5-1, and Gemini-3.1-Pro. Each model was given a standardized prompt (reproduced in Supplementary Text S1) that identifies the trial, defines success as meeting the registered primary endpoint and failure as not meeting it, and requests a binary verdict; web search was enabled for every model, and the model calls were made between 2 and 15 October 2026. The API resource used for each model and the web-search configuration are detailed in Supplementary Text S2.

For inClinico Agent, predictions were generated on 28 September 2026. Following standard practice for a deployed model, the inClinico models were first refit on all available data (the combined training and test sets). Because each endpoint model outputs a continuous PTI, we determined a decision threshold for each of the efficacy and safety/toxicity models by maximizing classification accuracy on the combined data; these thresholds were then applied to convert the PTI into the binary success/failure predictions reported for the 3,235 benchmark trials. The binary predictions of inClinico Agent and the five language models for all 3,235 trials are provided in Supplementary Table 1.

ClinicoBench will be scored only once its trials read out, under a prespecified analysis plan (Supplementary Text S2); we will release inClinico Agent’s verdicts alongside those of the five language models periodically as individual trials complete, so that the comparison accrues as genuine prospective evidence rather than retrospective fit.

## Results

### Trial characteristics

The modeling cohort comprised 32,097 trials with a resolved success or failure outcome: 28,265 in the training set (start year 2000–2020) and 3,832 in the test set (start year 2021 or later), of which 21,690 were routed to the efficacy model and 10,407 to the safety/toxicity model (Table 1). Overall, 56.2% of trials met their primary endpoint. The composition of the two endpoint groups differed in two notable ways. First, first-in-class programs were more common among safety/toxicity trials than among efficacy trials (14.2% versus 8.8% of trials). Second, the phase distributions differed: efficacy trials were concentrated in Phases II and III (47.0% and 31.6%), whereas safety/toxicity trials were concentrated in Phases I and II (56.5% and 23.6%).

**Table 1.** Characteristics of trials for training and testing. Values are the number of trials and the within-column percentage. (Note: Phase counts do not sum to the column total because the trial phase is missing for a small number of trials.)

| Characteristic | Train set | Test set | Combined set |
| --- | --- | --- | --- |
| **Efficacy and safety/toxicity trials** |  |  |  |
| Total trials | 28,265 (100.0%) | 3,832 (100.0%) | 32,097 (100.0%) |
| Trial outcome |  |  |  |
| Success | 15,898 (56.2%) | 2,126 (55.5%) | 18,024 (56.2%) |
| Failure | 12,367 (43.8%) | 1,706 (44.5%) | 14,073 (43.8%) |
| First-in-class status |  |  |  |
| First-in-class | 2,807 (9.9%) | 568 (14.8%) | 3,375 (10.5%) |
| Non-first-in-class | 25,458 (90.1%) | 3,264 (85.2%) | 28,722 (89.5%) |
| Trial phase |  |  |  |
| Phase I | 6,293 (22.3%) | 1,373 (35.8%) | 7,666 (23.9%) |
| Phase II | 11,236 (39.8%) | 1,420 (37.1%) | 12,656 (39.4%) |
| Phase III | 7,646 (27.1%) | 694 (18.1%) | 8,340 (26.0%) |
| Phase IV | 3,059 (10.8%) | 342 (8.9%) | 3,401 (10.6%) |
| **Efficacy trials** |  |  |  |
| Total trials | 19,343 (100.0%) | 2,347 (100.0%) | 21,690 (100.0%) |
| Trial outcome |  |  |  |
| Success | 10,714 (55.4%) | 1,364 (58.1%) | 12,078 (55.7%) |
| Failure | 8,629 (44.6%) | 983 (41.9%) | 9,612 (44.3%) |
| First-in-class status |  |  |  |
| First-in-class | 1,603 (8.3%) | 295 (12.6%) | 1,898 (8.8%) |
| Non-first-in-class | 17,740 (91.7%) | 2,052 (87.4%) | 19,792 (91.2%) |
| Trial phase |  |  |  |
| Phase I | 1,455 (7.5%) | 332 (14.1%) | 1,787 (8.2%) |
| Phase II | 9,075 (46.9%) | 1,126 (48.0%) | 10,201 (47.0%) |
| Phase III | 6,260 (32.4%) | 601 (25.6%) | 6,861 (31.6%) |
| Phase IV | 2,526 (13.1%) | 285 (12.1%) | 2,811 (13.0%) |
| **Safety/toxicity trials** |  |  |  |
| Total trials | 8,922 (100.0%) | 1,485 (100.0%) | 10,407 (100.0%) |
| Trial outcome |  |  |  |
| Success | 5,184 (58.1%) | 762 (51.3%) | 5,946 (57.1%) |
| Failure | 3,738 (41.9%) | 723 (48.7%) | 4,461 (42.9%) |
| First-in-class status |  |  |  |
| First-in-class | 1,204 (13.5%) | 273 (18.4%) | 1,477 (14.2%) |
| Non-first-in-class | 7,718 (86.5%) | 1,212 (81.6%) | 8,930 (85.8%) |
| Trial phase |  |  |  |
| Phase I | 4,838 (54.2%) | 1,041 (70.1%) | 5,879 (56.5%) |
| Phase II | 2,161 (24.2%) | 294 (19.8%) | 2,455 (23.6%) |
| Phase III | 1,386 (15.5%) | 93 (6.3%) | 1,479 (14.2%) |
| Phase IV | 533 (6.0%) | 57 (3.8%) | 590 (5.7%) |

Observed success rates varied systematically with these characteristics (Supplementary Table 2). First-in-class trials succeeded less often than non-first-in-class trials (43.2% versus 57.7% overall), consistent with the greater biological uncertainty of novel targets. Success rate also rose with development phase, with later-phase trials succeeding more often than earlier-phase trials (combined success of about 65% in Phase III and 63% in Phase IV, versus about 50–53% in Phases I and II). The magnitude of these success rates is broadly consistent with those reported previously for inClinico (Aliper et al. 2023) and with historical estimates of clinical trial success rates (Wong et al. 2019).

### Model selection

Before fixing the production models, we compared the four candidate frameworks — logistic regression, XGBoost, LightGBM, and CatBoost — for each endpoint family, considering both their cross-validation performance on the training set (Supplementary Table 3) and their performance on the test set (Figure 4, Supplementary Table 4). The three tree-ensemble frameworks clearly outperformed logistic regression: in both endpoint families, and under both criteria, the AUROC and AUPRC confidence intervals of the tree ensembles showed little or no overlap with those of logistic regression. Among the three tree ensembles, by contrast, performance was very similar — their AUROC and AUPRC point estimates differed by only about 0.01–0.02 and their 95% confidence intervals overlapped substantially — so no single tree ensemble is clearly superior. We nonetheless selected, for each endpoint, the framework with a slight overall edge: XGBoost for efficacy (test AUROC 0.828, 95% CI 0.812–0.845) and CatBoost for safety/toxicity (test AUROC 0.809, 95% CI 0.787–0.831). Because the final choice took the test-set performance into account, the reported test metrics of the selected models may be modestly optimistic; however, the tree ensembles differed so little on the test set — and were statistically indistinguishable under both the cross-validation and the test criteria — that the specific choice among them has little practical consequence, and any such optimism is minimal.

**Figure 4.**
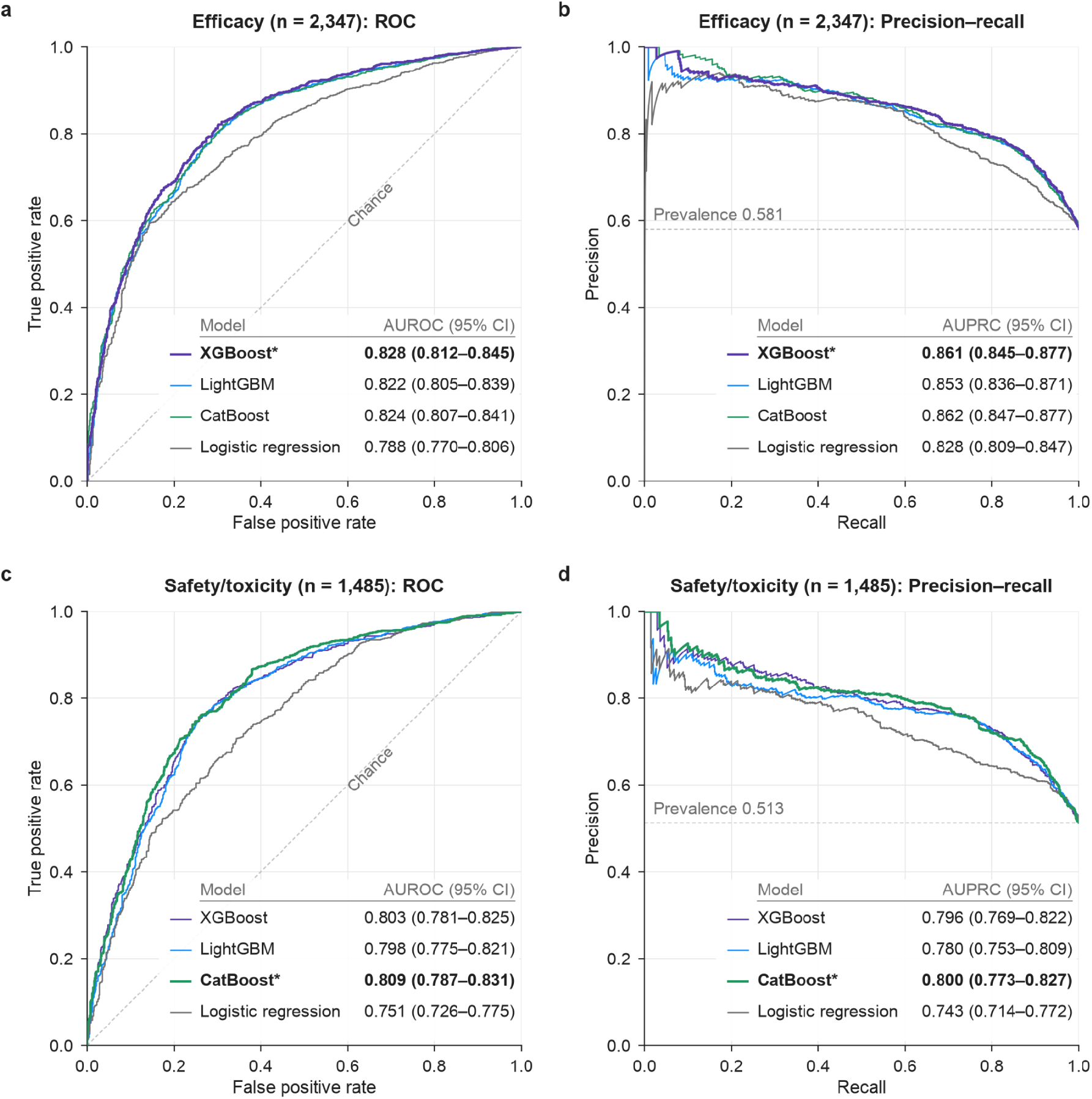
Candidate-model comparison on the test set. **a)** Receiver operating characteristic (ROC) curves for the efficacy endpoint (n = 2,347). **b)** Precision–recall curves for the efficacy endpoint. **c)** ROC curves for the safety/toxicity endpoint (n = 1,485). **d)** Precision–recall curves for the safety/toxicity endpoint. Each panel compares the four candidate frameworks: XGBoost, LightGBM, CatBoost, and logistic regression. The table in each panel gives the AUROC or AUPRC of each framework with its 95% confidence interval (CI). The asterisk, bold row, and thicker line mark the framework deployed for that endpoint. The dashed diagonal marks chance discrimination. The dashed horizontal line marks the prevalence of successful trials in the test set, which is the precision expected by chance. AUPRC, area under the precision–recall curve; AUROC, area under the ROC curve.

### Model performance

On the test set, the efficacy model performed slightly better than the safety/toxicity model (overall AUROC 0.828, 95% CI 0.812–0.845, versus 0.809, 95% CI 0.787–0.831; Figure 5). Within each endpoint family, discrimination was higher for non-first-in-class trials than for first-in-class trials (efficacy AUROC 0.828 versus 0.792; safety/toxicity 0.815 versus 0.790). The confidence intervals for the first-in-class subgroups are wide because those subgroups are small (295 efficacy and 273 safety/toxicity first-in-class trials in the test set); nevertheless, the gap is in the same direction for both endpoints, suggesting a real difficulty of predicting trial outcomes for novel targets. Performance also increased with development phase: although the per-phase confidence intervals overlap, the point estimates rise monotonically from Phase I to Phase III (for example, efficacy AUROC 0.796, 0.820, and 0.852 in Phases I, II, and III). Phase IV and the safety/toxicity Phase III subgroup contained relatively few trials, and their estimates are correspondingly noisier.

**Figure 5.**
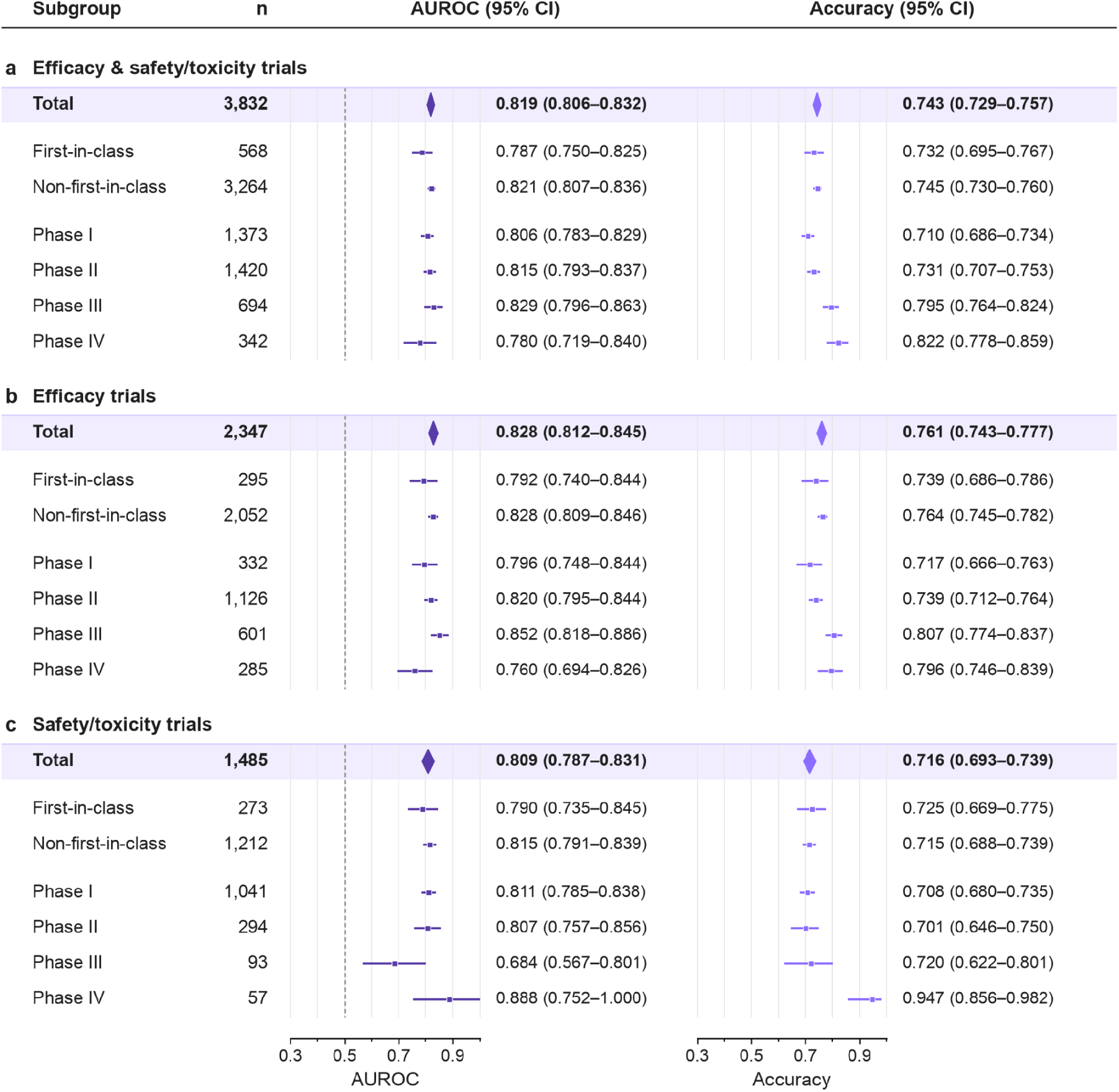
Endpoint-prediction performance on the test set, by cohort and subgroup. AUROC (left) and accuracy (right) for **a)** efficacy and safety/toxicity trials combined, **b)** efficacy trials, and **c)** safety/toxicity trials, shown overall and by first-in-class status and trial phase. Metrics for the combined cohort pool the trials of the two endpoint groups. Because the two endpoint groups are scored by separate models, the combined-cohort AUROC and AUPRC should be read as a summary across both models rather than as a single-model metric; the per-endpoint rows (b, c) are the primary results. Accuracy uses model-specific decision thresholds that maximized accuracy on the training-set cross-validation out-of-fold predictions. Squares mark point estimates and horizontal lines mark 95% confidence intervals (CIs). Diamonds mark cohort totals and span their 95% CIs. The n column gives the number of trials in each subgroup. (Note: Phase counts may not sum to the column total because the trial phase is missing for a small number of trials.)

Because the dataset is curated up to the second half of 2026, 2025 is the most recent year for which the corpus contains a complete and representative set of trials with resolved success or failure outcomes. We therefore additionally evaluated the models on trials completing in 2025 (Supplementary Figure 1). The patterns described above generally held: the efficacy model outperformed the safety/toxicity model, discrimination was higher for non-first-in-class than first-in-class trials, and performance tended to increase with phase. Overall performance in 2025 (combined AUROC 0.805, 95% CI 0.777–0.833) was close to performance on the entire test set (AUROC 0.819, 95% CI 0.806–0.832).

### ClinicoBench

Because the 3,235 ClinicoBench trials are ongoing, their true outcomes are not yet known, and it therefore remains unknown whether inClinico Agent or the large language models will ultimately prove more accurate. Two patterns are nonetheless already apparent (Figure 6). First, inClinico Agent predicted success for about half of the benchmark trials, close to the historical success rate observed in the test set, whereas each of the five language models predicted success far more often — for roughly three-quarters or more of the trials (Figure 6a). Second, chance-corrected agreement among the models was highly uneven (Figure 6b). We quantified agreement with Cohen’s kappa, which removes the agreement expected from each model’s own predicted-positive rate; raw percent agreement would otherwise be inflated simply because the language models predict success so frequently. The five language models agreed substantially with one another (pairwise kappa approximately 0.43–0.67), whereas inClinico Agent showed only slight, near-chance agreement with any of them (kappa approximately 0.12–0.18). inClinico Agent is therefore making materially different calls from the language models.

**Figure 6.**
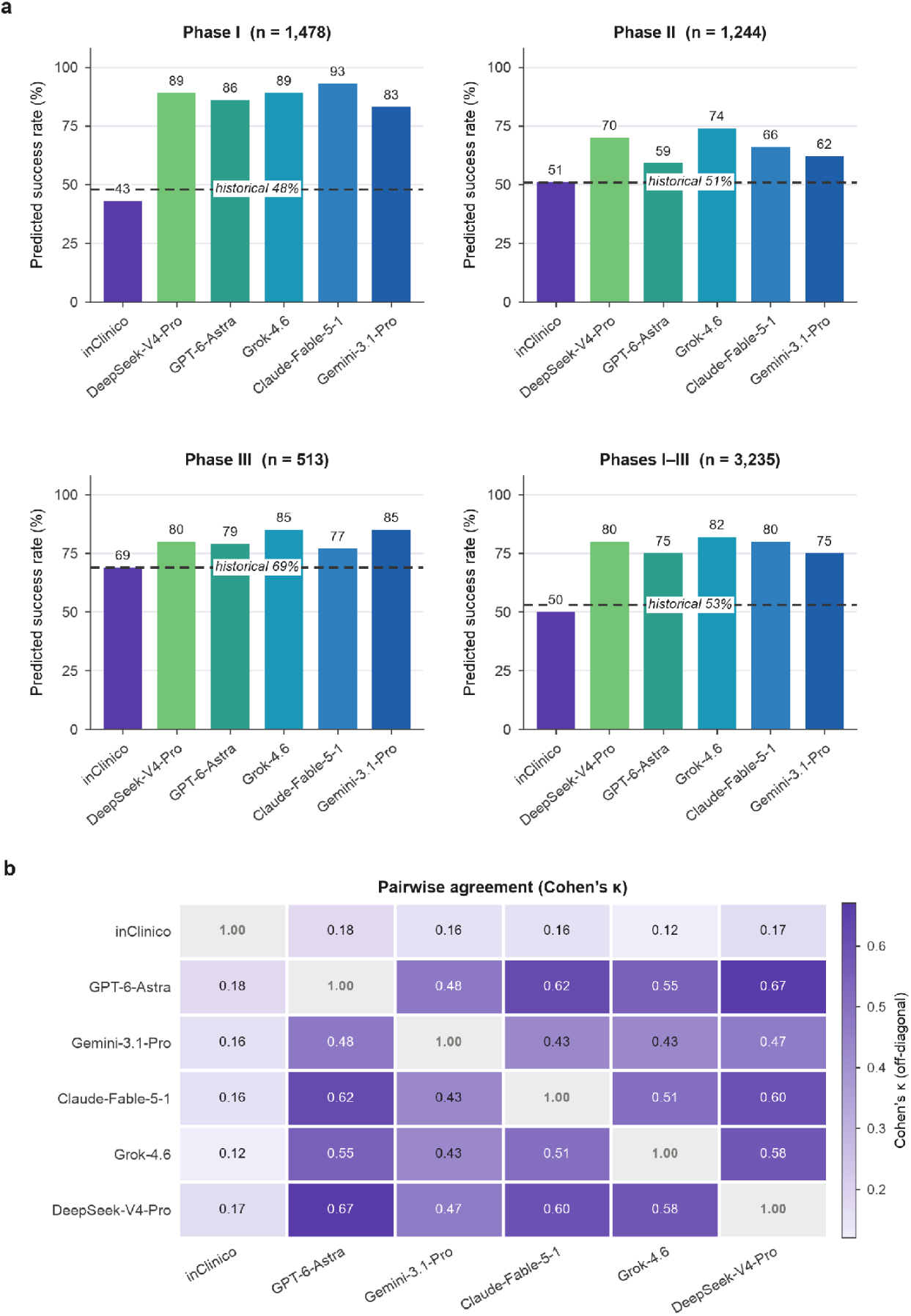
ClinicoBench predictions and inter-model agreement on 3,235 ongoing trials. a) Percentage of ClinicoBench trials predicted to succeed by inClinico Agent and five frontier LLMs, by trial phase (Phase I, II and III, and Phases I–III combined). The dashed horizontal line gives the observed success rate of trials of that phase in the test set. The n in each panel title is the number of ClinicoBench trials in that phase. b) Pairwise chance-corrected agreement between models, measured by Cohen’s κ on the binary success/failure verdicts. Cohen’s κ removes the agreement expected from each model’s own predicted-positive rate. Unlike raw percent agreement, it is therefore not inflated when two models both predict success frequently. It quantifies concordance between models and does not measure correctness. Darker cells indicate higher κ, and the diagonal is self-agreement (κ = 1).

Because the benchmark trials have not yet read out, ClinicoBench is a living, forward evaluation; we will release inClinico Agent’s verdicts alongside those of the five language models periodically over the following months as individual trials read out, so that the comparison accrues as genuine prospective evidence rather than retrospective fit.

## Discussion

We describe an endpoint-specific machine-learning model that predicts clinical trial success across all phases, along with the agentic AI system in which it is embedded. Three features of the modeling approach are worth emphasizing. First, by training and evaluating the efficacy and safety/toxicity endpoints separately, the model respects the distinct biology and distinct failure modes of the two questions, rather than collapsing them into a single notion of “success.” Second, the time-based separation of training and test data — training and tuning only on trials initiated before 2021, with time-blocked cross-validation inside the training set, and evaluating on trials initiated afterward — provides a realistic estimate of how the model will behave in deployment, where it must always predict the future from the past. Performance was similar on the 939 trials completing in 2025 and consistent across phases, indicating that the test-set estimates are stable rather than specific to a single slice of the data. Third, the model deliberately abstains from using any feature that would only become available after a trial is registered or under way. Such features can inflate apparent performance in a retrospective evaluation, but they would be unavailable — and would therefore degrade performance — in a genuinely prospective setting. This approach is intended to keep the reported performance representative of what the model will achieve on future trials.

The test set is challenging by construction. It contains a higher proportion of first-in-class programs (14.8% versus 9.9% in the training set) and of early-phase trials (Phase I rising from 22.3% to 35.8%) than the training data — precisely the trials that are harder to predict, as the subgroup results make clear. We deliberately applied no statistical adjustment to compensate for this distributional shift; thus, the numbers we report represent the model’s unadjusted performance on the real-world distribution of recent trials.

ClinicoBench addresses a different limitation of the retrospective testing. A retrospective AUROC, however carefully computed, is always open to the criticism that the labels already existed when the model was built; a benchmark of trials that have not yet read out is not. The original inClinico study did include a genuinely prospective evaluation, but on only 17 trials and restricted to Phase II (Aliper et al. 2023). An important contribution of ClinicoBench is to extend prospective evaluation to more than 3,000 ongoing trials spanning all phases and both endpoint families. Benchmarking against five frontier LLMs with web search enabled further situates the model against the strongest available general-purpose baselines. Finally, as these trials read out, a systematic analysis of the cases that inClinico Agent and the language models get wrong should yield actionable guidance both for improving the inClinico model and for understanding when a language model can be relied upon for this task. We will release ClinicoBench outcomes in stages as trials complete.

Beyond the predictive model, inClinico Agent is designed as a transparent, auditable analytical environment.

### Autonomous agent architecture

The system is powered by a dynamic multi-agent orchestration layer (Figure 1) that categorizes each objective, routes it to the appropriate reasoning or retrieval component, and executes end-to-end workflows, pairing frontier LLMs for contextual reasoning with deterministic engines for data retrieval, scoring, and visual reporting. It provides live visibility into reasoning steps, intermediate thinking, and tool calls, and maintains durable per-session logging so that multi-turn follow-up remains grounded in a preserved workspace.

### Data provenance and verified reporting

Each session operates within an isolated workspace that archives raw retrieval outputs, intermediate artifacts, and final deliverables. Evidence is weighed through a deterministic hierarchy that prioritizes primary literature (identified by PMID or DOI) and registry records over computed model signals, and citations are drawn strictly from tool execution. An automated verification layer checks that each claim is actually grounded in its cited source, rather than merely that a link resolves, which guards against unsubstantiated citations. Deliverables — including radar charts and competitive-landscape visualizations — are generated in Markdown, HTML, and PDF with interactive, stream-labeled references that remain faithful across export formats.

### Specialized clinical skills

The quantitative model is surfaced through an integrated suite of specialized skills that span the clinical-development workflow. These include head-to-head comparison of two competing programs; evaluation of existing trials across a cohort to produce a competitive leaderboard; scoring of a proposed trial prior to registration, benchmarked against comparable historical studies and accompanied by a mandatory literature-grounding pass; response-biomarker analysis on real transcriptomic datasets; drug-safety and regulatory assessment built on openFDA adverse-event, labeling, and approval records; clinical-trial-protocol drafting aligned to ICH standards and configured for multiple regulatory authorities; clinical-pharmacology strategy, including first-in-human dose derivation; and regulatory-submission support across the major dossier types and review modes. These skills form the Core Skills layer of the inClinico Agent architecture, where they are grouped into trial evaluation and trial preparation (Figure 1).

### Future directions

A weakness of the current model is its comparatively modest performance on first-in-class programs. Two directions seem promising for closing this gap. First, much of the signal for a novel target is relational — which pathways, diseases, and prior programs it connects to — and could be captured by a carefully curated, comprehensive, high-quality biomedical knowledge graph with explicitly time-stamped edges, coupled with graph-representation models; time-stamping is essential so that only knowledge available before a trial’s start informs its prediction, preserving the leakage-free design. Second, the drugs and targets themselves carry molecular structure that our current literature-derived scores capture only indirectly; embeddings from well-established molecular and protein encoders could add complementary, mechanism-aware features. We consider both realistic avenues to improve prediction, particularly for first-in-class trials.

## Conclusions

We have presented an endpoint-specific machine-learning model that predicts clinical trial success across all phases using only information available at the time of trial design, and the agentic AI system, inClinico Agent, that delivers it alongside a suite of specialized clinical skills. On a test set of trials initiated in 2021 or later, the efficacy and safety/toxicity models reached AUROCs of 0.828 and 0.809, and performance was stable on trials completing in 2025. Through ClinicoBench, we are now evaluating the model prospectively on more than 3,000 ongoing trials against five frontier large language models, with results to be released as the trials read out. By pairing leakage-free predictions with transparent, auditable reporting, inClinico Agent is designed to help clinical-development teams prioritize trials and anticipate their outcomes.

## Data Availability

The curated clinical trial database used to train the models in this study is not publicly available. The prospective ClinBench predictions generated in this study are provided in Supplementary Table 1.

## Supplementary Material

**Supplementary Figure 1.**
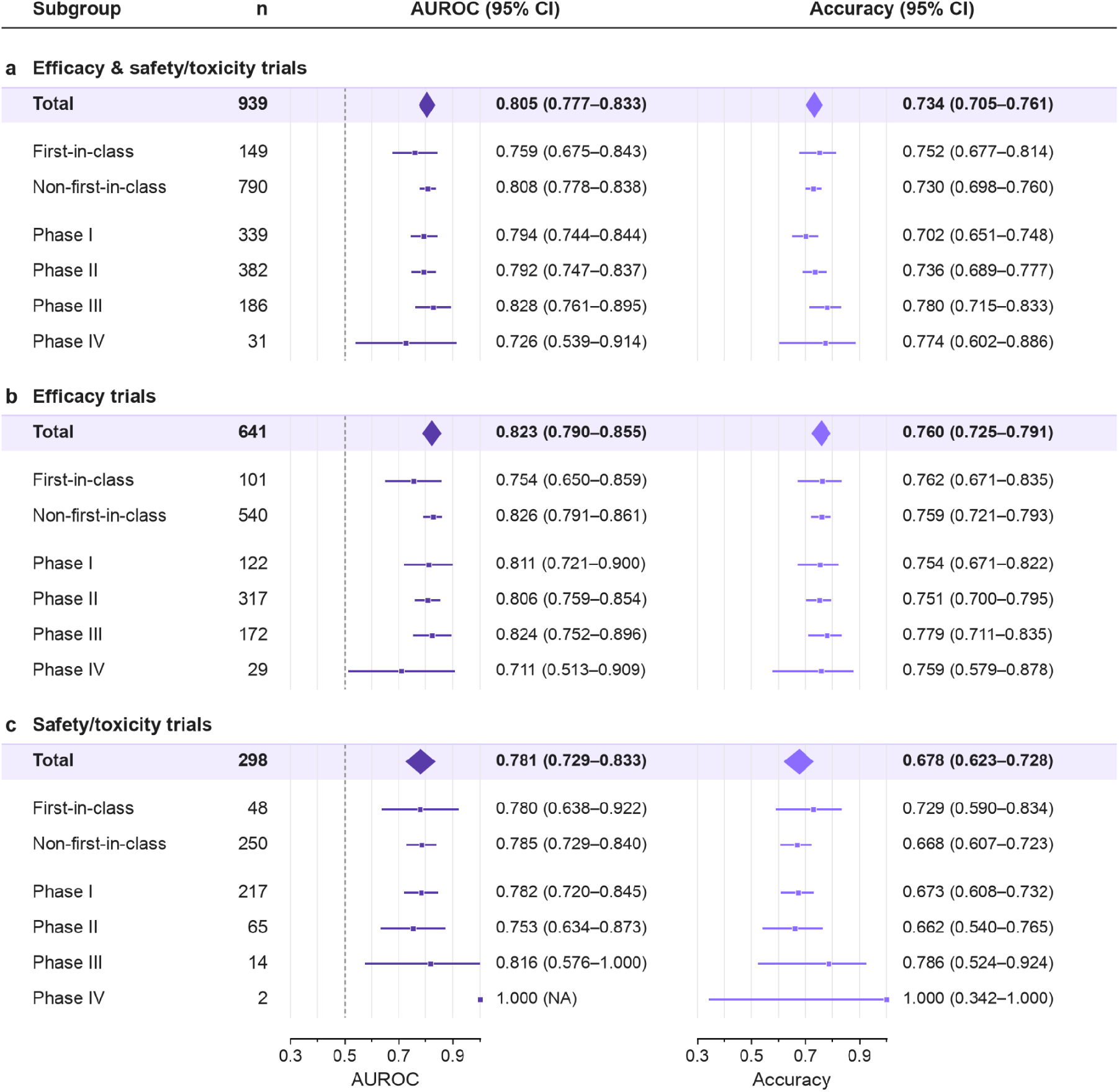
Endpoint-prediction performance on the test-set trials completing in 2025, by cohort and subgroup. Similar to Figure 5, except restricted to trials whose completion date falls in 2025. (Note: Because there are only two Phase IV safety/toxicity trials, the associated 95% confidence interval degenerates and is shown as NA. Phase counts may not sum to the column total because the trial phase is missing for a small number of trials.)

**Supplementary Table 1.** Binary ClinicoBench predictions. Provided separately as suppl_table_1.csv, this table lists the binary success/failure prediction of inClinico Agent and each of the five language models for all 3,235 benchmark trials, together with the trial’s registry identifier, phase, and each model’s prediction timestamp (28 September 2026 for inClinico Agent; the date of the call for each language model).

**Supplementary Table 2.**
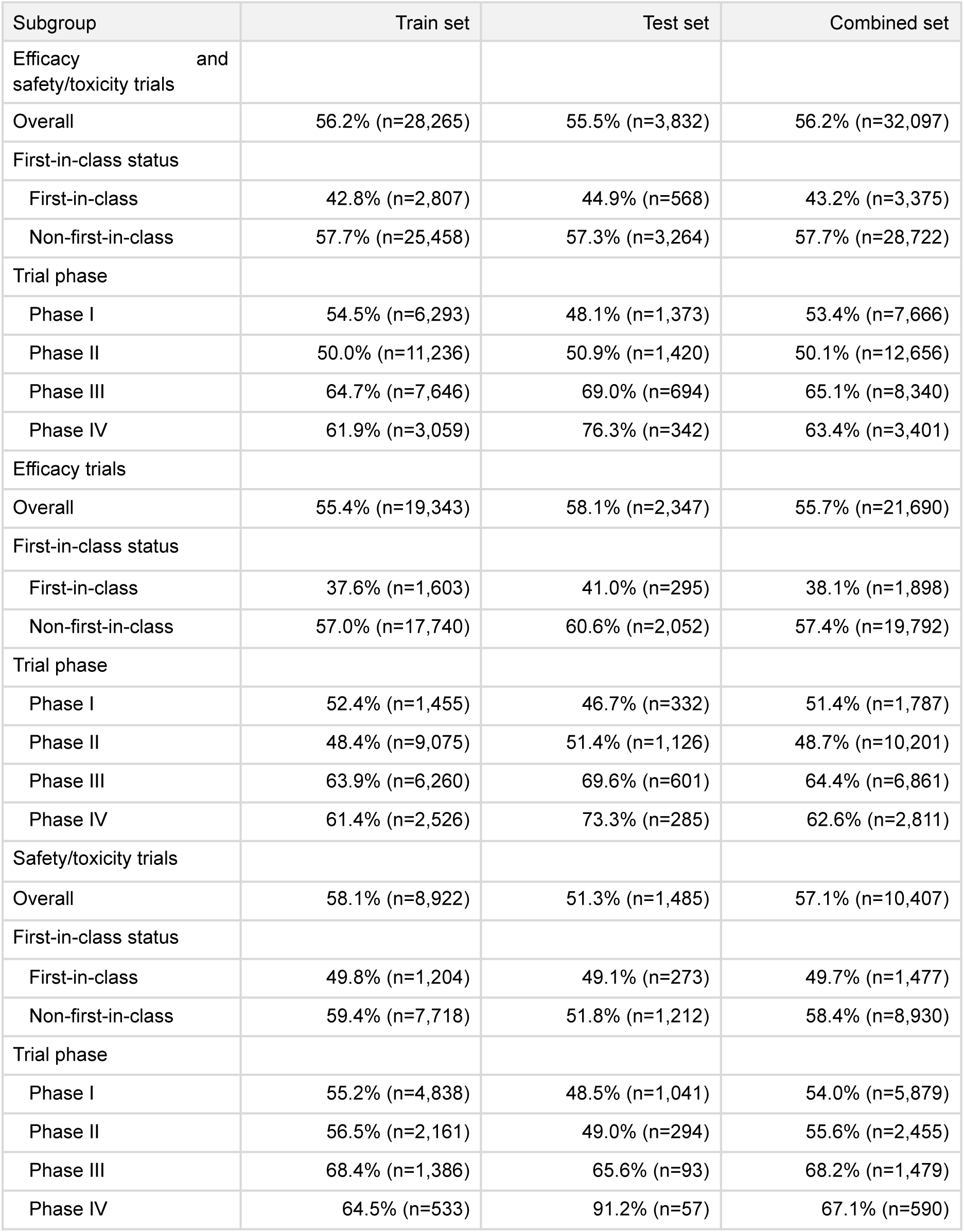
Percentage of trials that succeeded, by subgroup. Each cell shows the percentage of trials that met their primary endpoint, followed in parentheses by the number of trials in that subgroup (successes and failures combined), which is the denominator of the percentage. (Note: Phase counts may not sum to the column total because the trial phase is missing for a small number of trials.)

**Supplementary Table 3.** Cross-validation performance (AUROC and AUPRC with 95% confidence intervals) for every candidate model. Metrics are computed on the pooled out-of-fold predictions of the time-blocked cross-validation on the training set (trials with a start year of 2000–2020).

| Model | AUROC (95% CI) | AUPRC (95% CI) |
| --- | --- | --- |
| Efficacy trials (out-of-fold n = 16,115) |  |  |
| XGBoost | 0.780 (0.773–0.787) | 0.813 (0.806–0.821) |
| LightGBM | 0.780 (0.772–0.787) | 0.814 (0.807–0.821) |
| CatBoost | 0.782 (0.775–0.790) | 0.818 (0.811–0.825) |
| Logistic regression | 0.751 (0.744–0.759) | 0.794 (0.787–0.802) |
| Safety/toxicity trials (out-of-fold n = 7,435) |  |  |
| XGBoost | 0.742 (0.731–0.754) | 0.796 (0.785–0.807) |
| LightGBM | 0.742 (0.730–0.753) | 0.797 (0.785–0.807) |
| CatBoost | 0.747 (0.736–0.758) | 0.804 (0.794–0.815) |
| Logistic regression | 0.688 (0.676–0.700) | 0.760 (0.748–0.771) |

**Supplementary Table 4.** Extended performance metrics (with 95% confidence intervals) for every candidate model on the test set. AUROC and AUPRC are threshold-free; accuracy, balanced accuracy, F1, precision, and recall are computed at the decision threshold described in Methods (maximized on the training-set cross-validation out-of-fold predictions, never on the test set). * marks the model used in the deployed pipeline for that endpoint (XGBoost for efficacy, CatBoost for safety/toxicity).

| Model | Decision thresholds | AUROC | AUPRC | Accuracy | Balanced accuracy | F1 | Precision | Recall |
| --- | --- | --- | --- | --- | --- | --- | --- | --- |
| Efficacy trials (n = 2,347) |  |  |  |  |  |  |  |  |
| XGBoost* | 0.457 | 0.828<br>(0.812–0.845) | 0.861<br>(0.845–0.877) | 0.761<br>(0.743–0.777) | 0.739<br>(0.721–0.756) | 0.809<br>(0.797–0.822) | 0.753<br>(0.732–0.774) | 0.874<br>(0.855–0.890) |
| LightGBM | 0.495 | 0.822<br>(0.805–0.839) | 0.853<br>(0.836–0.871) | 0.762<br>(0.745–0.779) | 0.746<br>(0.728–0.764) | 0.805<br>(0.792–0.819) | 0.768<br>(0.746–0.789) | 0.846<br>(0.826–0.864) |
| CatBoost | 0.492 | 0.824<br>(0.807–0.841) | 0.862<br>(0.847–0.877) | 0.764<br>(0.747–0.781) | 0.748<br>(0.731–0.766) | 0.807<br>(0.793–0.821) | 0.771<br>(0.749–0.791) | 0.847<br>(0.827–0.865) |
| Logreg | 0.493 | 0.788<br>(0.770–0.806) | 0.828<br>(0.809–0.847) | 0.719<br>(0.700–0.737) | 0.698<br>(0.679–0.716) | 0.774<br>(0.760–0.788) | 0.727<br>(0.704–0.748) | 0.828<br>(0.807–0.847) |
| Safety/toxicity trials (n = 1,485) |  |  |  |  |  |  |  |  |
| XGBoost | 0.523 | 0.803<br>(0.781–0.825) | 0.796<br>(0.769–0.822) | 0.723<br>(0.699–0.745) | 0.719<br>(0.697–0.741) | 0.761<br>(0.743–0.778) | 0.682<br>(0.652–0.711) | 0.860<br>(0.833–0.882) |
| LightGBM | 0.485 | 0.798<br>(0.775–0.821) | 0.780<br>(0.753–0.809) | 0.715<br>(0.692–0.738) | 0.711<br>(0.689–0.732) | 0.761<br>(0.746–0.777) | 0.668<br>(0.638–0.696) | 0.886<br>(0.861–0.906) |
| CatBoost* | 0.482 | 0.809<br>(0.787–0.831) | 0.800<br>(0.773–0.827) | 0.716<br>(0.693–0.739) | 0.712<br>(0.690–0.733) | 0.765<br>(0.748–0.780) | 0.666<br>(0.637–0.694) | 0.898<br>(0.874–0.917) |
| Logreg | 0.493 | 0.751<br>(0.726–0.775) | 0.743<br>(0.714–0.772) | 0.661<br>(0.637–0.685) | 0.656<br>(0.634–0.677) | 0.725<br>(0.709–0.742) | 0.621<br>(0.592–0.650) | 0.871<br>(0.846–0.893) |

### Supplementary Text S1: ClinicoBench prompt

Each LLM received the following prompt for every benchmark trial, with the trial’s registry identifier substituted for {NCT_ID}:

Evaluate this active clinical trial {NCT_ID} (outcomes unreported). Analyze the trial details and predict its binary outcome: Success or Failure.

You must go to the trial’s page on clinicaltrials.gov (https://clinicaltrials.gov/study/{NCT_ID}) to check its study design, phase, status, and enrollment.

A trial is defined as successful if its primary endpoints are met. A trial is defined as a failure if its primary endpoints are not met.

Return ONLY valid JSON: {“success”:0 or 1,“confidence”:0.0–1.0,“reasoning”:“<=50 words”}

### Supplementary Text S2: ClinicoBench model access and prespecified analysis plan

**Model access and web search.** The five language models were accessed through a mix of cloud-hosted deployments and the vendors’ own APIs: GPT-6-Astra on Azure OpenAI and Claude-Fable-5-1 on Azure AI Foundry; Gemini-3.1-Pro on Google Cloud Vertex AI; and Grok-4.6 and DeepSeek-V4-Pro through their providers’ official APIs (xAI and DeepSeek, respectively). Web search was enabled as a provider-side server tool for every model, so that each could retrieve current public information about a trial at prediction time: a web_search tool on the Responses API for GPT and Grok, a web_search tool on the Anthropic Messages interface for Claude and DeepSeek, and Google Search grounding for Gemini. The prediction timestamp for each call is recorded alongside the per-trial predictions in Supplementary Table 1.

**Prespecified analysis plan.** ClinicoBench will be scored only once its trials read out. Because each model is asked for a binary success/failure verdict and the two outcome classes are not perfectly balanced, our primary metric will be balanced accuracy. The models differ in the kind of output they produce: a conventional binary classifier such as inClinico Agent emits a continuous score — a predicted probability, equivalently a log-odds — that can be rank-ordered to trace a receiver operating characteristic or precision–recall curve, whereas a large language model returns a discrete success/failure verdict together with a self-reported confidence rather than such an internal continuous score. For a supplementary, threshold-free comparison we will nonetheless map each language model’s output to a continuous score — the reported confidence when it predicts success, and one minus the confidence when it predicts failure — and compute AUROC and AUPRC from that score, noting that these self-reported confidences are not model-internal probabilities and may be poorly calibrated.

## Notes

### Competing Interest Statement

All authors are affiliated with Insilico Medicine: Insilico Medicine is a global clinical-stage commercial generative artificial intelligence company with several hundred patents, pending patent applications, and commercially available software.

### Summary of Updates

(1) We changed the name of the prospective benchmark set of clinical trials from ClinBench to ClinicoBench. (2) We reduced the number of trials for benchmarking from 3,311 to 3,235, excluding 76 trials that had readout released before the LLM call, which ended on 2026-10-15.

## References

Akiba, Takuya, Shotaro Sano, Toshihiko Yanase, Takeru Ohta, and Masanori Koyama. 2019. “Optuna: A Next-Generation Hyperparameter Optimization Framework.” Proceedings of the 25th ACM SIGKDD International Conference on Knowledge Discovery and Data Mining, 2623–31. 10.1145/3292500.3330701.

Aliper, Alex, Roman Kudrin, Daniil Polykovskiy, et al. 2023. “Prediction of Clinical Trials Outcomes Based on Target Choice and Clinical Trial Design with Multi-Modal Artificial Intelligence.” Clinical Pharmacology & Therapeutics 114 (5): 972–80. 10.1002/cpt.3008.

Chen, Jintai et al. 2025. “TrialBench: Multi-Modal AI-Ready Datasets for Clinical Trial Prediction.” Scientific Data, ahead of print. 10.1038/s41597-025-05680-8.

Chen, Tianqi, and Carlos Guestrin. 2016. “XGBoost: A Scalable Tree Boosting System.” Proceedings of the 22nd ACM SIGKDD International Conference on Knowledge Discovery and Data Mining, 785–94. 10.1145/2939672.2939785.

DeLong, Elizabeth R., David M. DeLong, and Daniel L. Clarke-Pearson. 1988. “Comparing the Areas Under Two or More Correlated Receiver Operating Characteristic Curves: A Nonparametric Approach.” Biometrics 44 (3): 837–45. 10.2307/2531595.

Efron, Bradley, and Robert J. Tibshirani. 1994. An Introduction to the Bootstrap. Chapman & Hall/CRC.

Fu, Tianfan, Kexin Huang, Cao Xiao, Lucas M. Glass, and Jimeng Sun. 2022. “HINT: Hierarchical Interaction Network for Clinical-Trial-Outcome Predictions.” Patterns 3 (4): 100445. 10.1016/j.patter.2022.100445.

Gao, Chufan, Jathurshan Pradeepkumar, Trisha Das, Shivashankar Thati, and Jimeng Sun. 2026. “A Large-Scale Database for Clinical Trial Outcomes and Features.” Nature Health 1: 428–36. 10.1038/s44360-026-00081-6.

Jin, Shuyi, Lu Chen, Hongru Ding, Meijie Wang, and Lun Yu. 2024. “Can Artificial Intelligence Predict Clinical Trial Outcomes?” arXiv:2411.17595. https://arxiv.org/abs/2411.17595.

Ke, Guolin, Qi Meng, Thomas Finley, et al. 2017. “LightGBM: A Highly Efficient Gradient Boosting Decision Tree.” Advances in Neural Information Processing Systems 30 (NeurIPS), 3146–54.

Lee, Jinhyuk, Wonjin Yoon, Sungdong Kim, et al. 2020. “BioBERT: A Pre-Trained Biomedical Language Representation Model for Biomedical Text Mining.” Bioinformatics 36 (4): 1234–40. 10.1093/bioinformatics/btz682.

Liu, Fengchen, Haoyu Wang, Jaemin Cho, Dan Roth, and Andrew W. Lo. 2025. “AutoCT: Automating Interpretable Clinical Trial Prediction with LLM Agents.” Proceedings of the 2025 Conference on Empirical Methods in Natural Language Processing (EMNLP). 10.18653/v1/2025.emnlp-main.1575.

Prokhorenkova, Liudmila, Gleb Gusev, Aleksandr Vorobev, Anna Veronika Dorogush, and Andrey Gulin. 2018. “CatBoost: Unbiased Boosting with Categorical Features.” Advances in Neural Information Processing Systems 31 (*NeurIPS*), 6638–48.

Wang, Jiacheng et al. 2026. CT Open: An Open-Access, Uncontaminated, Live Platform for the Open Challenge of Clinical Trial Outcome Prediction.

Wang, Zifeng, Cao Xiao, and Jimeng Sun. 2023. “SPOT: Sequential Predictive Modeling of Clinical Trial Outcome with Meta-Learning.” Proceedings of the 14th ACM International Conference on Bioinformatics, Computational Biology, and Health Informatics (ACM-BCB). 10.1145/3584371.3613001.

Wilson, Edwin B. 1927. “Probable Inference, the Law of Succession, and Statistical Inference.” Journal of the American Statistical Association 22 (158): 209–12. 10.1080/01621459.1927.10502953.

Wong, Chi Heem, Kien Wei Siah, and Andrew W. Lo. 2019. “Estimation of Clinical Trial Success Rates and Related Parameters.” Biostatistics 20 (2): 273–86. 10.1093/biostatistics/kxx069.

Yue, Ling, Sixue Xing, Jintai Chen, and Tianfan Fu. 2024. “ClinicalAgent: Clinical Trial Multi-Agent System with Large Language Model-Based Reasoning.” Proceedings of the 15th ACM International Conference on Bioinformatics, Computational Biology, and Health Informatics (ACM-BCB). 10.1145/3698587.3701359.

Zheng, Wenhao et al. 2025. “LIFTED: Multimodal Clinical Trial Outcome Prediction via Large Language Models and Mixture-of-Experts.” Findings of the Association for Computational Linguistics: EMNLP 2025. 10.18653/v1/2025.findings-emnlp.396.

